# OVERVIEW OF ETHICS COMMITTEE REGISTRATION AND RE-REGISTRATION WITH THE DEPARTMENT OF HEALTH RESEARCH IN INDIA

**DOI:** 10.1101/2023.04.19.23288784

**Authors:** Yash Kamath, Yashashri Shetty, Ankita Kulkarni

## Abstract

The Government of India requires all ethics committees (EC) to register themselves with the Department of Health Research (DHR) and the study aimed to analyze the same. Since post-graduation mandates research – necessitating the presence of EC in the institution – we aimed to analyze the status of registrations along with the presence of ECs in National Medical Council(NMC) and Dental Council of India(DCI)-permitted colleges and also to study zone-wise differences in the same. We also studied the impact of COVID-19 on the registration process. The status of ECs was obtained from the DHR website. ECs information was extracted [location, date & type of Approval]. The search terms “College” and “Hospital” were used to locate Institutional ECs (IEC). A list of Medical and Dental colleges was obtained from the NMC and DCI Websites with their location information. Of 931 IECs, 103(11.06%) had final, 728(78.2%) had provisional approval while 100(10.74%) ECs registration had expired. The median delay to file for provisional approval was 857 (IQR=407) days. The expired ECs functioning without registration for 90 (IQR=131) days. 440 of 931 (47.3%) ECs got registered during the COVID-19 period. The IEC to Medical Colleges ratio shows a significant zone-wise difference (Kruskal-Wallis, p=0.019) with the Northern Zone having the highest mean ratio of 0.90(SD=0.63). It is vital to improve DHR compliance and the EC/College Ratio for appropriate ethical governance and improve international acceptance of Indian Research.

## INTRODUCTION

Prior to 19 March 2019, all Ethics Committees (ECs) in India conducting biomedical and health research were required to register themselves with the Central Drugs Standards Control Organisation(CDSCO). However, with the implementation of the New Drugs and Clinical Trial Regulations -2019(NDCTR-19)[1], the ECs were required to register themselves with the National Ethics Committee Registry for Biomedical and Health Research(NECRBHR)[2], which is governed by the Department of Health Research(DHR), Government of India.

The ECs are required to apply for registration along with supporting documents citing the constitution, experience of members, function and documentation of protocols reviewed by the EC. The DHR provides a provisional registration to the ECs applying for the first time. This registration is effective for two years, following which the EC must apply for final registration with the DHR. Once a review of the projects sanctioned by the EC during the two-year period is completed, they are granted a final registration for a period of five years. Under the earlier regulations, 1260 ECs had registered themselves with the CDSCO, as per Das et al.(2019)[3]. We undertook this study to assess the current numbers and status of registrations with DHR, and to analyse the impact of the COVID-19 pandemic on the process. We also further assessed the zone-wise distribution of ECs and checked if ECs were proportionate to the Postgraduate(PG) Medical and Dental Colleges in India.

## METHODS

We sought exemption from ethical review(EC/OA-124/2022) under the clause “research on public anonymized non-identifiable data” of the ICMR guidelines for exempt review. Once the approval was obtained, we began the process of data extraction from the Department of Health Research(DHR)[4] and the National Medical Commission(NMC), and Dental Council of India(DCI) websites. In India, Medical and Dental courses are regulated by the NMC[5] and DCI[6], respectively. July 15, 2022 was taken as the cutoff date for data extraction. All the data available at the time on the above-mentioned websites was downloaded. Being a government regulatory body, it is assumed that all the colleges(medical and dental) functioning in India would be registered with the NMC & DCI.

The current status of Ethics Committees was obtained from the DHR website. The data we retrieved included the location information (associated institute, address and state), date of approval by DHR and the type of approval (Final, Provisional, Expired). The list of ECs were then screened by institute name to select only those associated with government or private medical or dental colleges. These were the institute-affiliated ECs (IECs). A list of colleges offering postgraduate and superspeciality courses, and also approved by the NMC, was obtained from its Website, creating a list of 304 colleges, with their location data and Management (Government, Trust, Society, Private). A similar process was followed for the DCI Website, which created a list of 303 colleges with their institute name and state. We considered the COVID-19 pandemic period from March 20, 2020, to August 1, 2021, when lockdown restrictions were eased.[7]

### Inclusion Criteria

All ECs mentioned on the DHR website under the Final, Provisional and Expired registration categories were included as part of the data extraction and analysis. Similarly, all NMC and DCI permitted Government and Private colleges were extracted from the respective website.

### Variables

The existing variables from the extracted data such as state names and affiliated institutes were used to create new variables such as the number of ECs and IECs per state, as well as the number of medical and dental colleges per state. These were used to calculate the EC to medical and dental college ratio. The number of Ethics committees in a state was divided by the total number of medical colleges in the state to create the EC /Medical College ratio; similarly, the number of Institutional Ethics Committees in the state affiliated with the medical and dental colleges of the same state was used to create IEC/Medical College and IEC/Dental College Ratios respectively. States were clubbed into zones (North: Himachal Pradesh, Punjab, Uttarakhand, Uttar Pradesh and Haryana. East: Bihar, Orissa, Jharkhand, and West Bengal; West: Rajasthan, Gujarat, Goa and Maharashtra; South: Andhra Pradesh, Karnataka, Kerala and Tamil Nadu; Central: Madhya Pradesh and Chhattisgarh; North East: Assam, Sikkim, Nagaland, Meghalaya, Manipur, Mizoram, Tripura and Arunachal Pradesh) and similar EC and college numbers were computed for each zone.

### Statistical Analysis

The data after extraction was compiled in a Microsoft Office Excel 2016 spreadsheet. The data was analyzed for descriptives (means, medians) using JASP 16.0 software and choropleth maps were created using Datawrapper.

## RESULTS

A total of 931 ECs were extracted from the DHR website. Furthermore, lists of 304 medical colleges and 303 dental colleges were obtained from NMC and DCI websites respectively. There were a total of 242 IECs (186 were Medical IECs - attached to medical colleges and 56 were Dental IECs, attached to dental colleges). *(Figure 1)*

**Figure 1:**
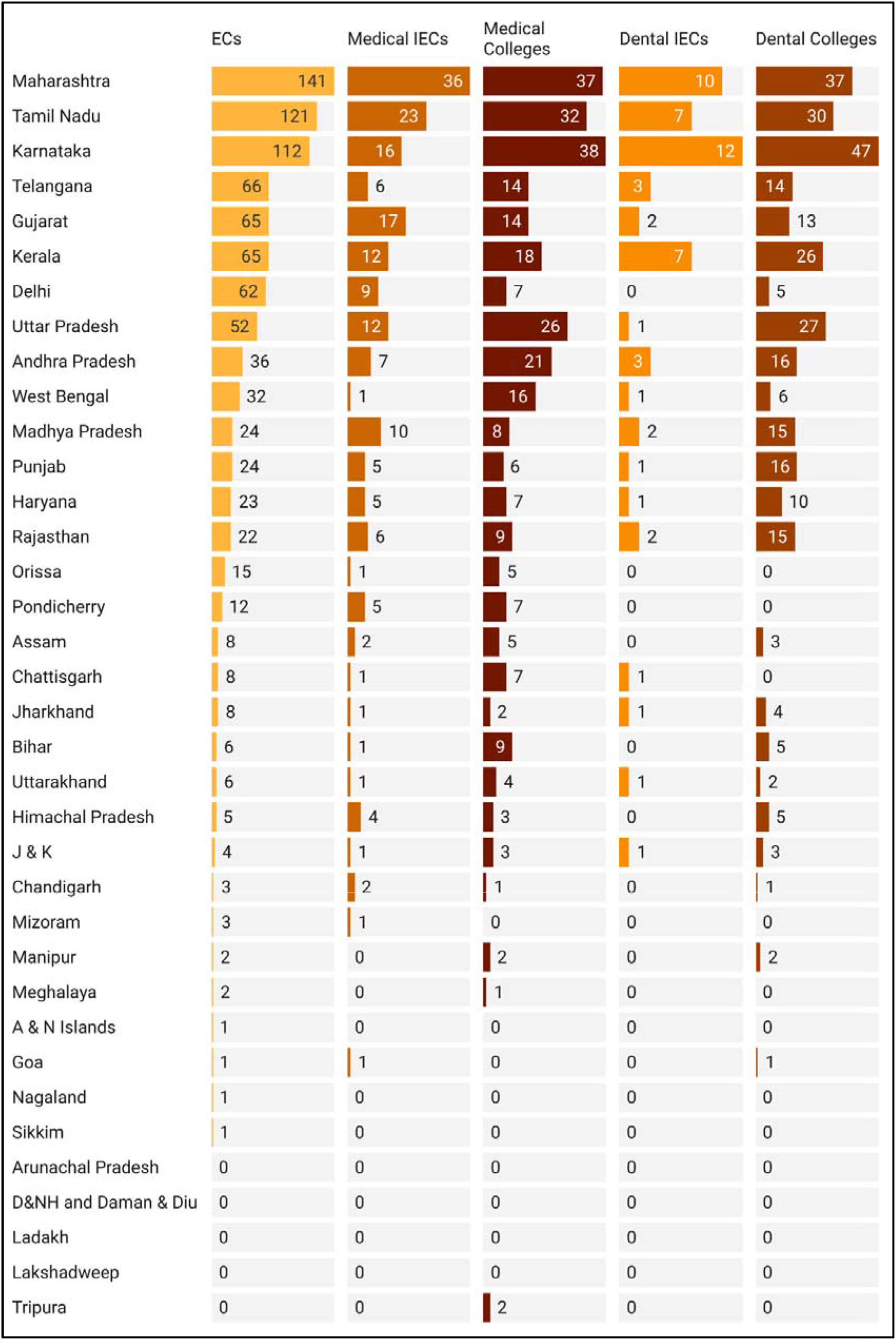
Statewise Distribution of ECs and IECs in India (EC = Ethics Committtees, IEC = Institutional Ethics Committees)(J&K = Jammu and Kashmir, A&N = Andaman and Nicobar, D&NH = Dadra and Nagar Haveli)

Of 931 ECs, 103(11.06%) had final approval and 728(78.2%) had provisional approval. 100(10.74%) ECs’ registration had expired at the time of access. The median delay in filing for provisional approval was 857 (IQR=407) days. The expired ECs were functioning without registration for a median of 90 (IQR=131) days. Among the 242 IECs, 33(13.64%) had final, 181(74.79%) had provisional and 28(11.57%) had expired approvals.

440 of 931 (47.3%) ECs got registered during the COVID-19 period. There was no significant difference in mean monthly registrations during and outside of the pandemic period, by the Mann-Whitney U-test (p=0.094). *(Figure 2)*

**Figure 2:**
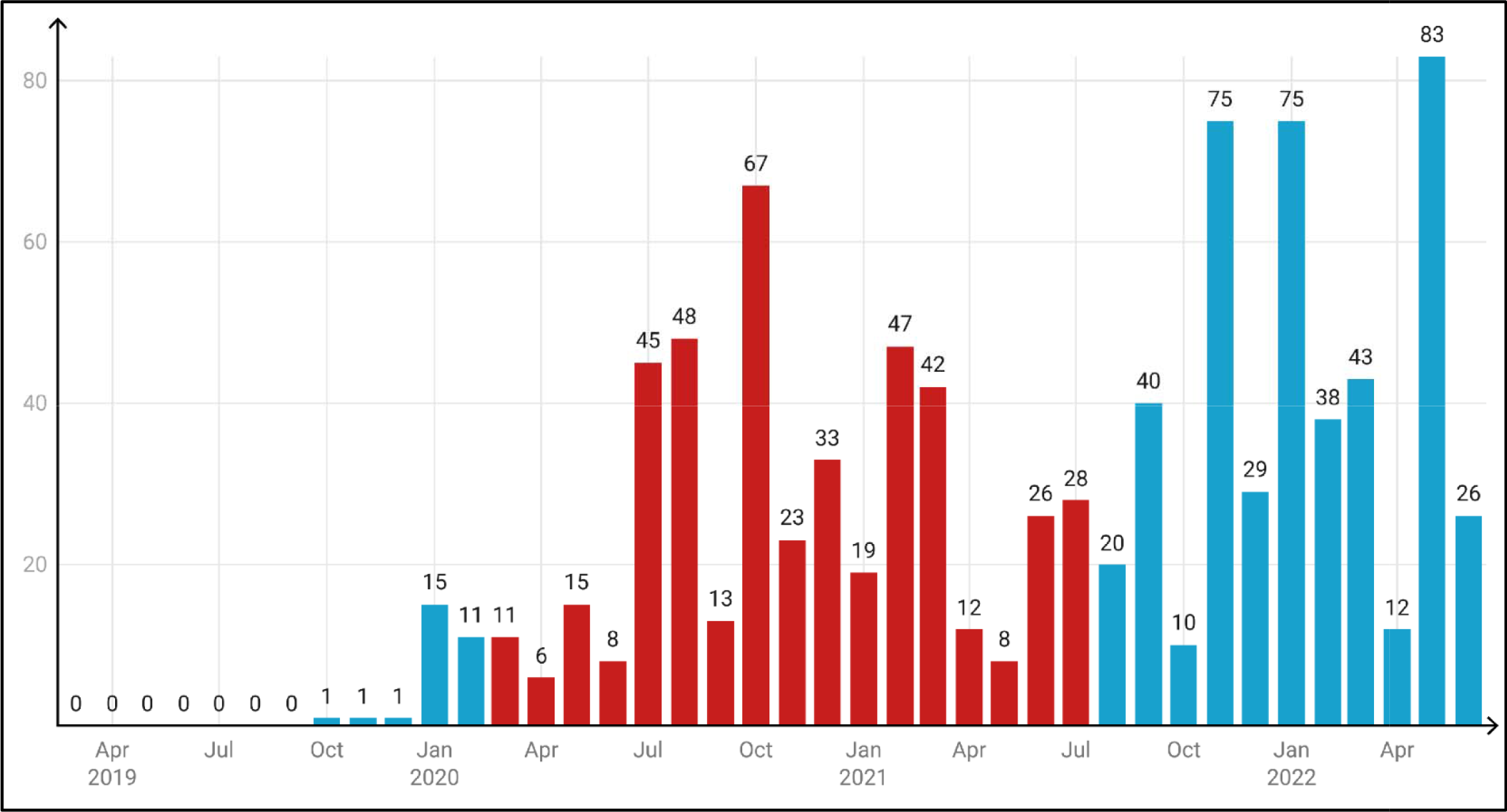
Ethics Committees arranged by Month and Year of DHR Registration. (COVID-19 Period registrations are marked in Red colour, Y Axis represents number of ethics committees registered during the month)

There was no significant zone-wise difference in the EC to Medical Colleges Ratio (Kruskal-Wallis, p=0.144); however, the Medical IEC to Medical College ratio shows a significant zone-wise difference (Kruskal-Wallis, p=0.019)*(Figures 3 & 4)* with Northern Zone having the highest mean ratio 0.90 ± 0.63 *(Table 1)*. The Dental IEC to Dental Colleges Ratio showed no significant zone-wise difference (Kruskal-Wallis, p=0.137).

**Table 1:** Zone-wise IEC and EC to College Ratios (IEC = Institutional Ethics Committees, EC = Ethics Committees)

| Zone | EC/Medical College Ratio | Medical IEC/Medical College Ratio | Dental IEC/Dental College Ratio |
| --- | --- | --- | --- |
| CENTRAL | 1.91 ± 0.81 | 0.53 ± 0.5 | 0.17 ± 0.23 |
| EAST | 2.42 ± 1.42 | 0.22 ± 0.2 | 0.1 ± 0.12 |
| NORTH | 3.07 ± 2.65 | 0.9 ± 0.63 | 0.08 ± 0.12 |
| NORTHEAST* | 0(1.15) | 0 | 0 ± 0 |
| SOUTH | 2.31 ± 1.75 | 0.41 ± 0.29 | 0.14 ± 0.12 |
| WEST | 2.11 ± 2.46 | 0.55 ± 0.64 | 0.11 ± 0.13 |
| **INDIA** | **3.06** | **0.61** | **0.18** |
| *\*for the Northeast Zone, Median (Interquartile Range) is reported.* |  |  |  |

**Figure 3:**
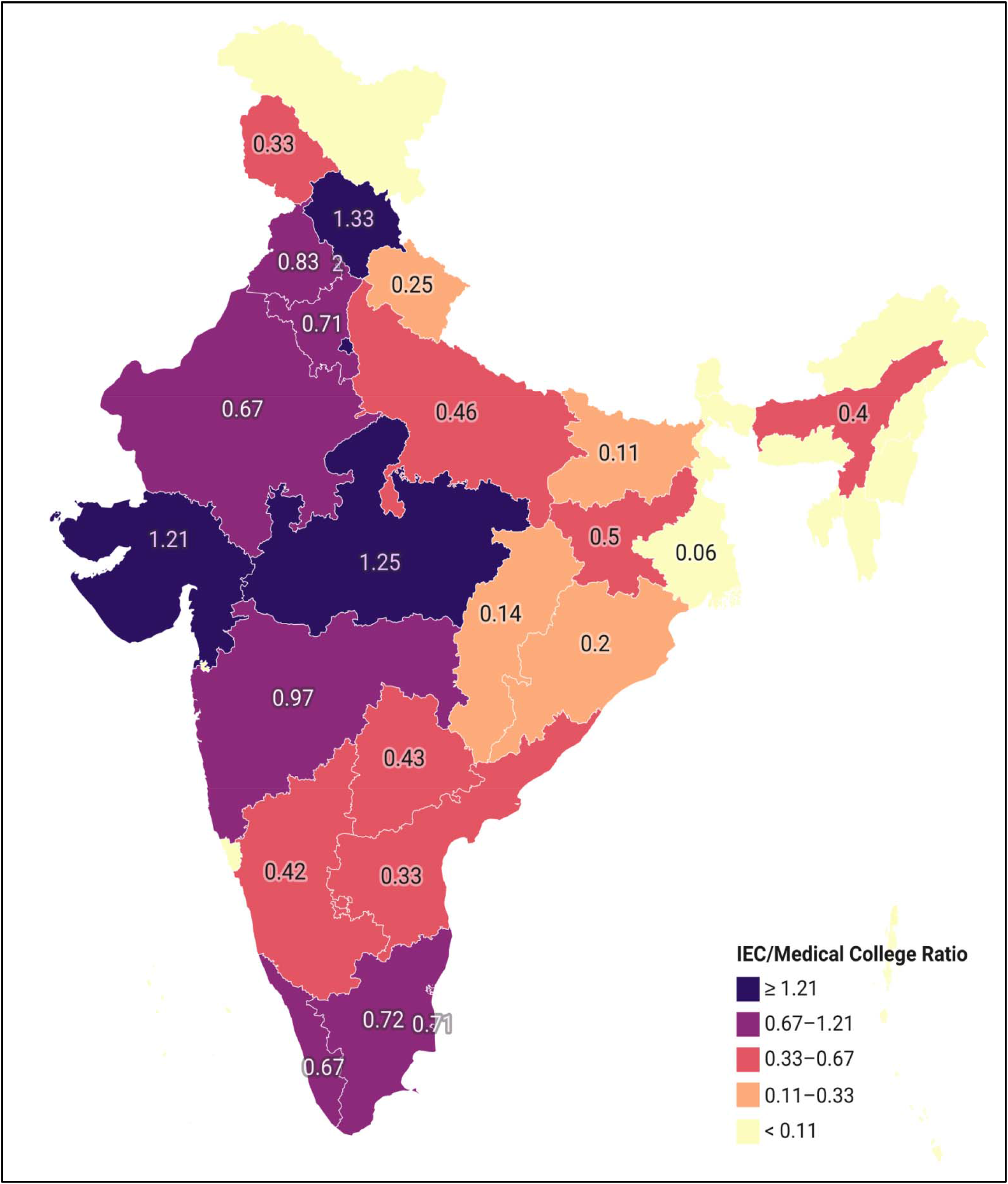
Medical EC/Medical College Ratio Statewise Heatmap (EC = Ethics Committee)

**Figure 4:**
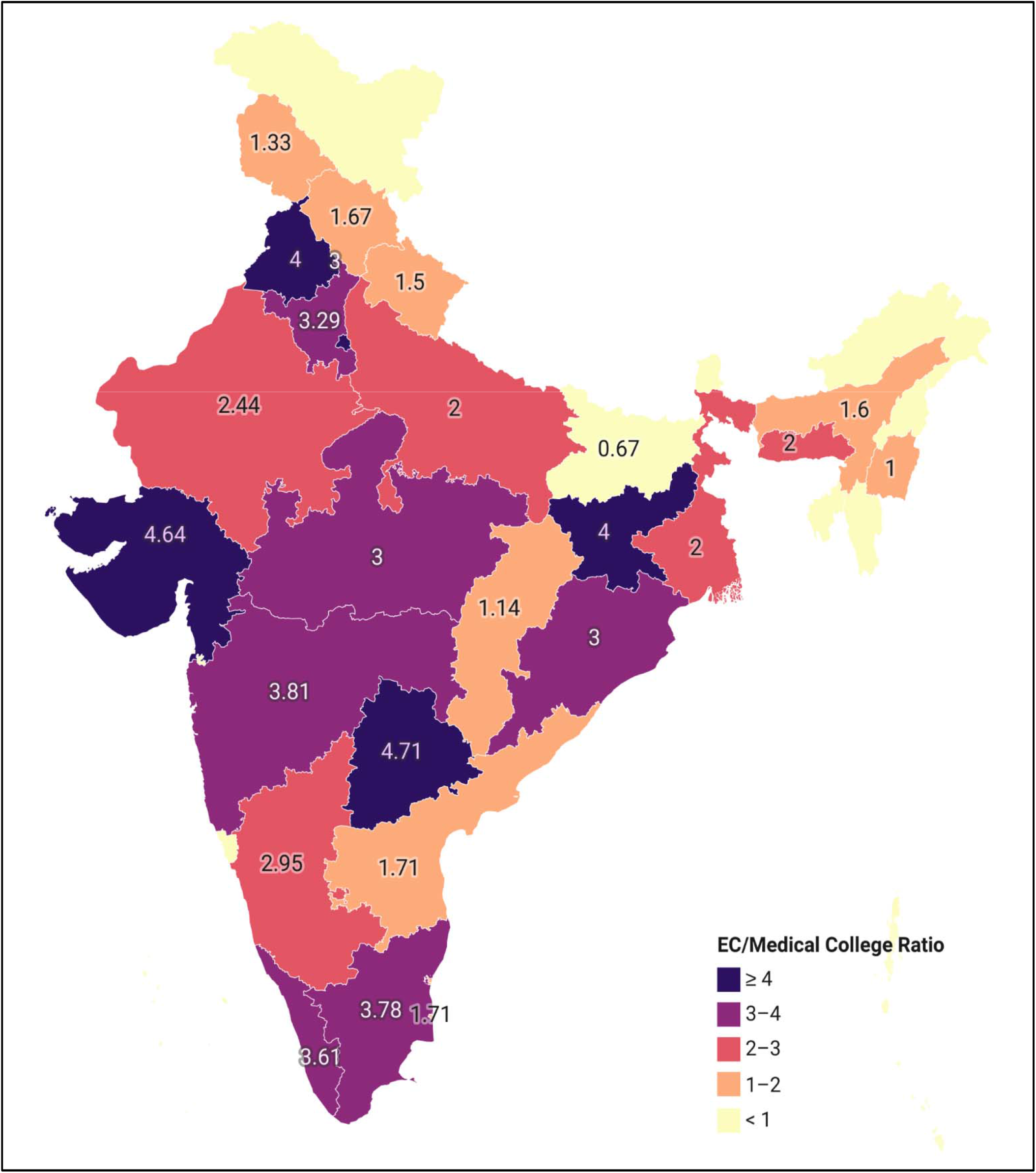
Medical IEC/Medical College Ratio Statewise Heatmap (IEC = Institutional Ethics Committee)

## DISCUSSION

The DHR collects information about the composition of the EC, the curriculum vitae of all its members, its Standard Operating Procedures (SOPs), Guidelines followed by the EC, copies of previously reviewed protocols, EC meetings, and all related administrative records. It also collects records relating to Serious Adverse Event reporting, medical management, and compensation provided, besides other minor points of information. Upon the receipt of this information from the EC, the DHR reviews it and decides on granting or rejecting the application for provisional approval. If it is granted, the approval lasts for a period of 2 years before it is required to be renewed. If ECs do not comply with the mandate, they may be suspended or their registration may be canceled. This is an indirect way of oversight on the ethics committees functioning by the Government of India, as an in-person audit of the Ethics committee is restricted by the little manpower working at the centers, money, and time.

We found that at least 10.7% of the ECs and 11.6% of IECs in DHR records, currently have an expired license. However, under the NMC Postgraduate Medical Education Regulations[8], thesis writing is part of the PG curriculum. Since thesis projects are essentially research in one form or another – they necessitate the presence of an ethics committee in the college offering a PG course to ensure ethical oversight which is mandatory as per ICMR guidelines 2017[9]. It does not however feature in the requirements to set up a new course or college under the NMC regulations.[10]

It was found that there was a delay in registration following the notification by the Government of India, ranging from 450 to 1264 days (interquartile range). This could be due to various factors, namely: institutional delay by the ethics committee, delay at the level of the regulatory body (DHR) or delay due to malfunctions in the submission platform. However, there is lack of evidence to support any of these reasonings behind the delay that clearly exists. It is our speculation that a combination of these factors is to blame.

The COVID-19 pandemic did not affect the registration process for the ECs (Figure 1) simply because the process is completely online and can be completed remotely without physical checks. We believe having the online mode available to the ECs through the pandemic ensured that no bottlenecks arose in the process and reviews and approvals were granted with ease. The ICMR guidelines issued during the pandemic in 2020[11], while emphasizing the need for DHR registration of ECs also stated the importance of COVID-19-related research. It is possible that these factors in conjunction might have further driven the ECs to register, so as to facilitate research in their institutes.

The IEC/Medical College and IEC/Dental College ratios suggest a trend that each college affiliated to the NMC or the DCI does not have its own Ethics Committee to assess and permit research projects. However, it is important to bear in mind that as compared to the 931 ECs recorded by us, the CDSCO website shows a total of 1400 EC registrations as of July 15, 2022[12] and Das et.al[3] have counted 1260 ECs in 2019. The higher number in CDSC0 registration can be due to the inclusion of not only medical schools but all the private/ government hospitals and research institutes. As CDSCO is a regulatory body, compliance to it is compulsory or else you will not be able to conduct regulatory studies, so the numbers are more. This implies that ECs are yet to completely adopt the DHR registration process. Even assuming that all ECs do complete DHR registration, the ratios which stand at 0.61 and 0.18(Table 1) are unlikely to reach the ideal of 1. But now as this biomedical and health research is part of the guidance in the NDCTR 2019 regulations from March 19, 2019, but even then compliance is not seen at medical colleges.

In the zone-wise comparison, we found that north and western zones performed better in IEC/Medical college ratios – we propose that this is due to a larger number of medical colleges in South India. However, this brings up the question of whether an Ethics Committee is necessary for the functioning of medical colleges – especially considering the colleges under question are providing postgraduate courses. In the north zones, Delhi has a major contribution with a large number of medical colleges as well as ethics committees(Figure 2). However, in overall performance, Maharashtra leads all the states, with 141 ECs and an IEC/Medical College ratio of 0.97. Southern and western states of India perform better in terms of EC numbers, but they do not match the medical or dental colleges in the state, which explains why the north and central regions perform better in these aspects.

It is vital to expand the scope and scrutiny of research done in India that the registration process for ECs be completed as swiftly as possible. Regular checks to clear ECs with an expired EC and make the steps to apply simpler may aid in achieving complete coverage. While the current global perception questions the quality of research done in India[13-16], we must strive to improve the pain points, starting with the most fundamental aspect – ethical review and accountability.

## CONCLUSION

There is a need to improve compliance of the existing ECs to the norms prescribed by DHR and Indian regulators. It is also vital to improve the EC to College Ratio, so as to ensure that ethical research is being undertaken in all institutes of India.

## Data Availability

All data produced in the present study are available upon reasonable request to the authors

## Author Contributions

The authors confirm their contribution to the paper as follows:

1. study conception and design: YK, YS
2. data collection: YK, YS, AK
3. analysis and interpretation of results: YK, YS, AK
4. draft manuscript preparation: YK, YS, AK

All authors reviewed the results and approved the final version of the manuscript..

## Acknowledgements

NA

